# Clinical prediction model performance in differentiating septic arthritis from transient synovitis: A multi-center study

**DOI:** 10.1101/2024.02.10.24302532

**Authors:** Xin Qiu, Han-Sheng Deng, Gen Tang, Yu-Xi Su, Xiao-Liang Chen, Yao-Xi Liu, Jing-Chun Li, Xin-Wu Wu, Jia-Chao Guo, Fei Jiang, Qi-Ru Su, Sheng-Ping Tang, Zhu Xiong, Chinese Multi-center Pediatric Orthopedic Study Group (CMPOS)

## Abstract

**Objective:** Differentiating septic arthritis from transient synovitis in children is challenging. This study aimed to determine the diagnostic value for distinguishing these two conditions and to develop an effective clinical prediction model based on multi-center clinical data.

**Methods:** We retrospectively analyzed data of children aged under 18 years who were hospitalized in eight specialized children’s hospitals in China from 2013 to 2021. To ensure the prediction model’s reliability, we established three clinical prediction models.

**Results:** This study collected data of 819 children from 8 tertiary children’s hospitals, including 265 patients with septic arthritis and 554 patients with transient synovitis.

We established three clinical prediction models. For septic hip arthritis, a retrospective study based on six clinical predictors was a history of prodromal respiratory tract infection (HRTI), temperature>37.5 °C, ESR>20 mm/h, CRP>10 mg/L, red blood cell distribution width (RDW)>50%, and WBC>11×109 /L. When these six factors were present, the probability of septic hip arthritis was 99.99%.

For septic knee arthritis, a retrospective study based on three clinical predictors, the predictors were ESR>20 mm/h, CRP>10 mg/L, and absolute monocyte count (AMONO)>0.74×109/L. When these three factors were present, the probability of having septic knee arthritis was 94.68%. For septic arthritis (septic hip arthritis or septic knee arthritis), a retrospective study based on six clinical predictors, the predictors were male children, history of HRTI), temperature>37.5 °C, ESR>20 mm/hr, PC > 407 × 10^9^/L and CRP>10 mg/L. When these six factors were present, the probability of septic arthritis was 99.65%.

**Conclusion:** This study used multi-center clinical data to construct a new clinical prediction model for children with septic arthritis. In addition we identified new clinical predictors such as sex, history of HRTI, RDW, PC and AMONO.

**Translational potential:** A clinical prediction model, built on multi-center data, is capable of effectively making high-precision predictions for septic arthritis. Furthermore, based on the microbial characteristics of septic arthritis in children, we aim to develop diagnostic kits that can accurately and quickly detect infections caused by pathogens such as bacteria.

## Introduction

Osteoarticular pain in children is an orthopedic emergency whose initial clinical presentation may pose a diagnostic challenge to the orthopedic surgeon, pediatrician, emergency room physician, or primary care physician [1]. Doctors usually differentiate between Legg-Perthes disease, slipped epiphysis of the femoral head, and fractures by collecting the patient’s medical history, conducting a physical examination, and performing radiography. They also consider septic arthritis and transient synovitis as the two most probable causes. However, these two medical conditions have similar clinical manifestations in the early stages: spontaneous and progressive joint pain, inability to bear weight on the affected limb, fever, and irritability. In addition, they require different treatments and have different outcomes. Septic arthritis is treated with antibiotics and surgical drainage, while transient synovitis is usually self-limiting and mainly managed symptomatically. Septic arthritis can lead to complications, such as osteonecrosis, growth arrest, and sepsis, whereas transient synovitis rarely has serious sequelae [2–4]. Therefore, early and accurate diagnosis of septic arthritis is crucial, and laboratory tests (such as ESR, WBC, and CRP levels) can sometimes be helpful. However, no definitive indicators can completely distinguish between the two conditions; therefore, clinical prediction models based on multiple factors may aid in diagnosis.

In 1999, Kocher et al. reported a clinical prediction model algorithm based on a retrospective study of four clinical predictors and prospectively validated it in 2004. The predictors were fever ≥38.5 °C, incapacity of the affected limb, weight-bearing, WBC >12×109 /L, and ESR ≥40 mm/h; when all predictors were positive, the probability of septic arthritis was 99.6% [1]. However, when Luhmann et al. retrospectively applied the same criteria, the predicted probability of septic arthritis was only 59% when all four clinical predictors were positive [5]. In 2006, Caird et al. used elevated CRP levels ≥20 mg/L as the fifth predictor, with a 98% probability of predicting septic arthritis [6].

Considering the differences in the accuracy of the aforementioned prediction models, we are currently unable to confirm the practicality of the clinical prediction models developed by each institution. Effective clinical prediction models may be developed in multi-center studies. Therefore, this study collected data through a review. Data on septic arthritis and transient synovitis in children from eight specialized children’s hospitals in China were collected to develop a clinical prediction algorithm model for septic arthritis and transient synovitis in Chinese children.

## Materials and methods

We retrospectively analyzed data of 819 children aged under 18 years who were hospitalized in eight specialized children’s hospitals in China from 2013 to 2021 and were diagnosed with septic arthritis or transient synovitis. Of them, 265 had septic arthritis (147 hip joints and 118 knee joints), and 554 had transient synovitis (487 hip joints and 67 knee joints).

We defined septic arthritis as a positive culture from a hip or knee aspirate or a positive blood culture from a hip or knee aspirate under high power with large numbers of white blood cells and no other identified source of infection[1, 5, 6], or as a clinical diagnosis made by the clinician at the first visit based on the child’s medical history (duration of symptoms, recent antibiotic treatment and reasons for treatment, fever and weight-bearing status), clinical symptoms, physical examination, and auxiliary examination results.

Transient synovitis was diagnosed in children with negative cultures, complete resolution of symptoms during follow-up, and no other confirmed joint-related disease[1, 5, 6], or as a clinical diagnosis is made by the clinician at the first visit based on the child’s medical history, clinical symptoms, physical examination, and auxiliary examinations.

The exclusion criteria are as follows: 1) osteomyelitis; 2) evidence of invasive bacterial infections in other sites, such as meningitis or cellulitis; 3) history of significant chronic or recurrent arthritis, rheumatism, bone or joint disease, or immunosuppressive disease, including malignancy, renal failure, inflammatory bowel disease, or any disease treated with immunosuppressive or anti-inflammatory drugs; and 4) purulent infections of the bones and joints caused by trauma, surgery, or iatrogenic causes.

Patients’ data included age, sex, initial hospital admission temperature, history of preceding respiratory tract infection, history of preceding strenuous exercise, duration of surgery, and name of surgery. Laboratory examinations included 36 indicators, such as ESR, WBC, CRP, PC, hematocrit, four coagulation parameters, and RDW. The etiological results included the results of pathogenic bacteria cultured from blood, affected joints, and surgical secretions. We defined fever as the first axillary body temperature greater than 37.5 °C on admission [7]. We defined a history of prodromal respiratory infection as the presence of an infection within the week before the initial evaluation. Further, we defined a history of prodromal strenuous exercise as the presence of strenuous exercise in the week before the initial evaluation.

The common sites of bone and joint infection in children are the hip and knee joints [8]. Therefore, to ensure the prediction model’s reliability, we established three clinical prediction models. The first model was for septic hip arthritis and transient hip synovitis, the second model was for septic knee arthritis and transient knee synovitis, and the third model was for septic arthritis and transient synovitis in either joint. We merged all the data of the hip and knee joints into one data set to build the third model [9].

Statistical methods: We used single-factor logistic regression analysis to screen candidate variables. We included indicators with p<0.2 in single-factor logistic regression analysis as candidate indicators. For the selected candidate indicators, we used different cutoff values to construct a single-factor logistic regression and used the cutoff value with the largest odd ratio (OR) as the cutoff value of the indicator. Backward selection stepwise multiple logistic regression was used to determine clinical predictors. We determined significance using the likelihood ratio chi-square test. Regression model fit was estimated using the Hosmer‒Lemeshow goodness-of-fit test, and calibration curves were used to assess model performance. We plotted the receiver operating characteristic curves (ROC) to evaluate the multivariable predictor panel in identifying septic arthritis diagnoses. Performance, sensitivity, and specificity were calculated using standard formulas. Furthermore, adjusted odds ratios and 95% confidence intervals were derived using the maximum likelihood method, and a nomogram was constructed to rapidly predict a patient’s probability of developing septic arthritis and to estimate the risk of septic arthritis of the hip for each combination. We performed statistical analyses using R software (version 4.3.1; R Foundation for Statistical Computing, Vienna, Austria.).

## Results

### Demographic information

A total of 819 children aged under 18 years who were evaluated in 8 tertiary pediatric hospitals in China from 2013 to 2021 were included, including 562 males and 257 females. Of them, 265 patients were finally diagnosed with septic arthritis (147 hip joints and 118 knee joints) and 554 had transient synovitis (487 hip joints and 67 knee joints). The average age of children with septic arthritis was 5.55±7.16 years, and the average age of children with transient synovitis was 4.8±3.2 years. Among the 265 patients with septic arthritis, 179 underwent surgical treatment, including 141 cases of incision and drainage + (Vacuum Sealing Draina, VSD) negative pressure drainage of the affected joints and 38 cases of joint lavage and drainage assisted by arthroscopy. In the blood and joint aspirate cultures of 265 children with septic arthritis, the most common causative organism was Staphylococcus aureus (59%), including 8 methicillin-resistant Staphylococcus aureus (MRSA) (12%), followed by Streptococcus pyogenes (10%), Streptococcus pneumoniae (7%), Staphylococcus epidermidis (3%), Klebsiella pneumoniae (3%), Haemophilus influenzae (3%), and other genera (15%).

### Univariate logistic regression analysis of septic hip arthritis and transient hip synovitis

Univariate logistic regression analysis showed significant differences (P<0.01) between septic hip arthritis and transient hip synovitis in the following variables: ESR, peripheral blood erythrocyte count, hemoglobin, CRP, RDW, history of preceding respiratory tract infection, AMONO, hematocrit, eosinophil percentage, lymphocyte percentage, WBC, initial hospital admission temperature, absolute neutrophil count (ANEC), platelet count (PC), platelet crit, mean corpuscular hemoglobin concentration, mean corpuscular volume, absolute eosinophil count, male sex, platelet distribution width, neutrophil percentage, age, and absolute lymphocyte count (Table 1).

### Univariate logistic regression analysis of septic knee arthritis and transient knee synovitis

Univariate logistic regression analysis revealed significant differences (P<0.01) between septic knee arthritis and transient knee synovitis in terms of ESR, CRP, AMONO, eosinophil percentage, initial hospital admission temperature, hemoglobin, WBC, serum erythrocyte count, PC, hematocrit, and platelet crit (Table 2).

### Univariate logistic regression analysis of septic arthritis (septic hip arthritis or septic knee arthritis) and transient synovitis (transient hip synovitis or transient knee synovitis)

Univariate logistic regression analysis showed significant differences (P<0.01) between septic arthritis (septic hip arthritis or septic knee arthritis) and transient synovitis(transient hip synovitis or transient knee synovitis) in the following variables: ESR, CRP, AMONO, eosinophil percentage, initial hospital admission temperature, hemoglobin, WBC, serum red blood cell count, PC, hematocrit, platelet crit, ANEC, absolute eosinophil count, mean corpuscular hemoglobin concentration, lymphocyte percentage, absolute lymphocyte count, history of preceding respiratory tract infection, neutrophil percentage, age, RDW, mean corpuscular volume, platelet distribution width, and male sex (Table 3).

### Multivariate analysis of septic hip arthritis and transient hip synovitis

We identified six independent multivariable predictors for distinguishing septic hip arthritis from transient hip synovitis as follows: having ESR>20 mm/h, CRP>10 mg/L, RDW>50%, WBC> 11×10^9^/L, temperature>37.5 ℃, and HRTI (Table 4). The prediction model based on these six predictors had a sensitivity of 88% and a specificity of 86 %.

### Multivariate analysis of septic knee arthritis and transient knee synovitis

We identified three independent multivariable predictors for distinguishing septic knee arthritis from transient knee synovitis as follows: having ESR>20 mm/h, CRP>10 mg/L, and AMONO>0.74×10^9^ /L (Table 5). The prediction model based on these three predictors had a sensitivity of 72% and a specificity of 88%.

### Multivariate analysis of septic arthritis (septic hip arthritis or septic knee arthritis) and transient synovitis (transient hip synovitis or transient knee synovitis)

We identified six independent multivariable predictors for distinguishing septic arthritis (septic hip arthritis or septic knee arthritis) from transient synovitis (transient hip synovitis or transient knee synovitis) as follows: having ESR >20 mm/h, CRP>10 mg/L, temperature>37.5 ℃, male sex, PC>407×10^9^ /L, and HRTI (Table 6). The prediction model based on these six predictors had a sensitivity of 84% and a specificity of 92%.

### Prediction of septic hip arthritis

We constructed an algorithm to determine the predicted probability of septic hip arthritis based on six independent multivariable predictors (Figure 1a). For example, if a child had HRTI, temperature>37.5 ℃, ESR>20 mm/h, CRP>10 mg/L, RDW>50%, and WBC>11×10^9^ /L, the probability of having septic hip arthritis was 99.99%. The area under the ROC curve was 0.93, indicating the excellent diagnostic performance of this set of six multivariate predictors in identifying septic hip arthritis (Figure 1b). The calibration plot showed good prediction accuracy between the actual and predicted probabilities (Figure 1c).

**Figure 1.**
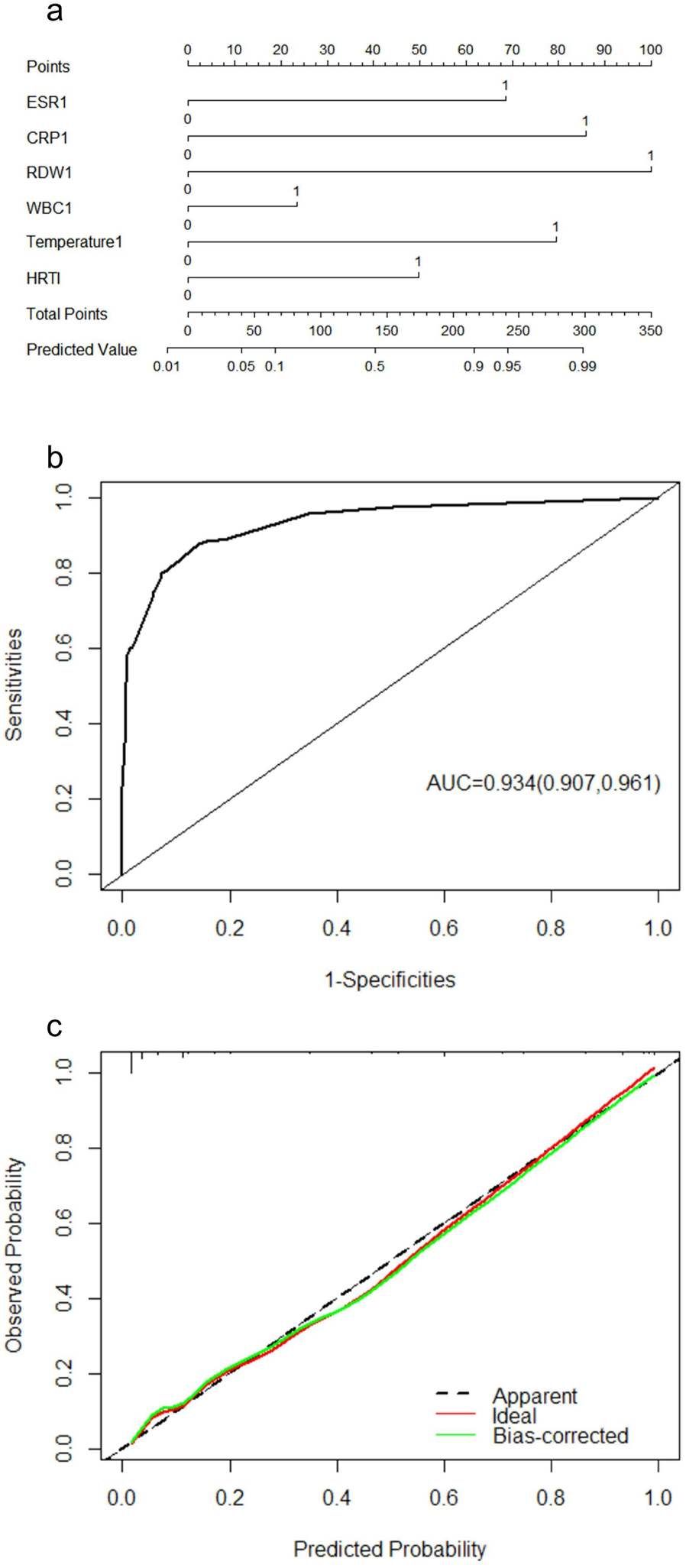
a. Nomogram of the clinical prediction model for septic hip arthritis. b. ROC curve of clinical prediction model for septic hip arthritis c. Calibration curve of the clinical prediction model for septic hip arthritis. The actual curve in the figure, the calibration curve, and the ideal curve have a high degree of fit.

### Prediction of septic knee arthritis

We constructed an algorithm to determine the predicted probability of septic knee arthritis based on three independent multivariable predictors (Figure 2a). For example, if a child’s blood test showed ESR>20 mm/h, CRP>10 mg/L, and AMONO>0.74×10^9^/L, the probability of having septic knee arthritis was 94.68%. The area under the ROC curve was 0.87, indicating the excellent diagnostic performance of this set of three multivariate predictors in identifying septic knee arthritis (Figure 2b). The calibration plot showed good prediction accuracy between the actual and predicted probabilities (Figure 2c).

**Figure 2.**
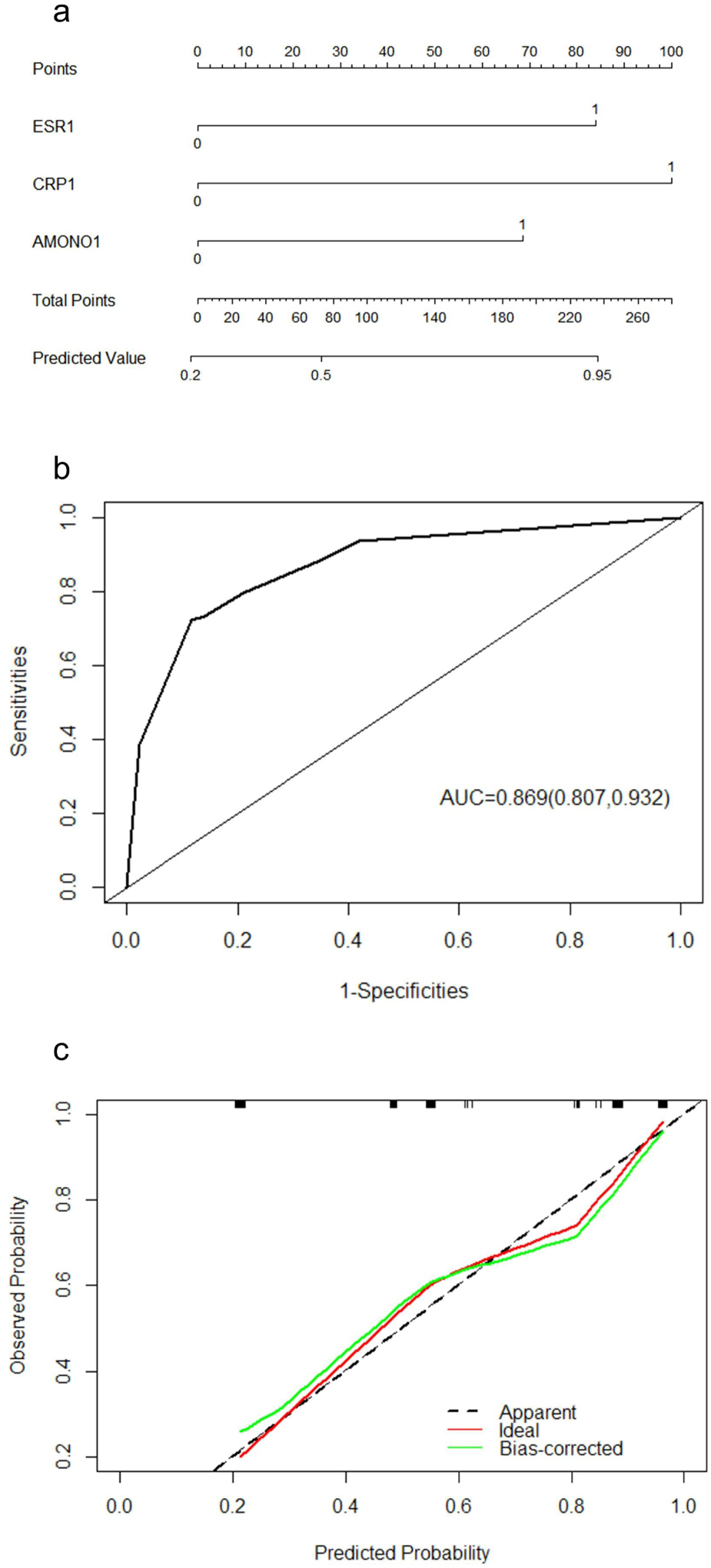
a. Nomogram of clinical prediction model for septic knee arthritis b. ROC curve of clinical prediction model for septic knee arthritis c. Calibration curve of the clinical prediction model for septic knee arthritis. The actual curve in the figure, the calibration curve, and the ideal curve have a high degree of fit.

### Prediction of septic arthritis (septic hip arthritis or septic knee arthritis)

We constructed an algorithm to determine the predicted probability of septic arthritis (septic hip or septic knee arthritis) based on six independent multivariate predictors (Figure 3a). For example, if a male child had HRTI, temperature>37.5 ℃, ESR>20 mm/h, CRP>10 mg/L, and PC>407×10^9^/L, the probability of septic arthritis was 99.65%. The area under the ROC curve was 0.94, demonstrating the excellent diagnostic performance of this set of six multivariate predictors in identifying septic arthritis (septic hip arthritis or septic knee arthritis) (Figure 3b). The calibration plot showed good prediction accuracy between the actual and predicted probabilities (Figure 3c).

**Figure 3.**
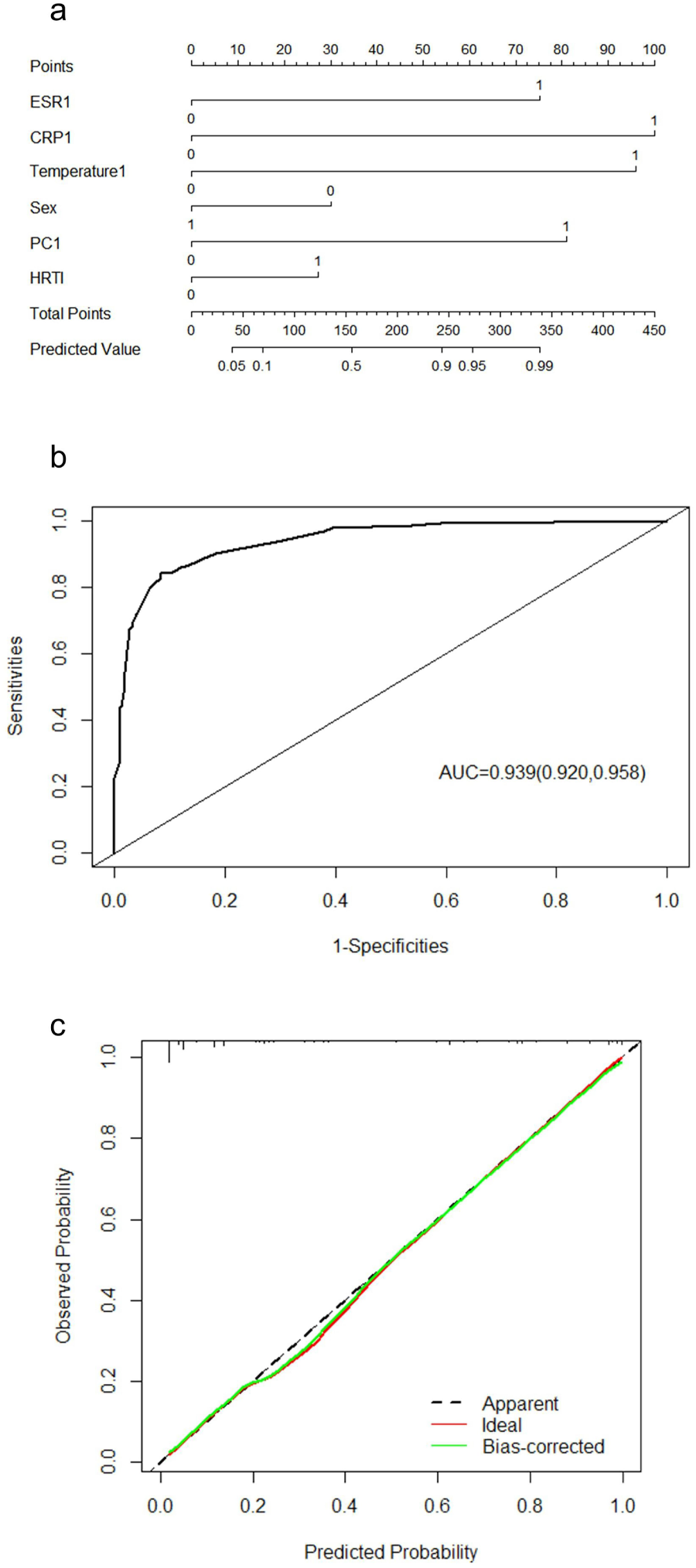
a. Nomogram of clinical prediction model for septic arthritis (septic hip arthritis or septic knee arthritis) b. ROC curve of the clinical prediction model for septic arthritis (septic hip arthritis or septic knee arthritis) c. Calibration curve of the clinical prediction model for septic arthritis (septic hip arthritis or septic knee arthritis). The actual curve in the figure, the calibration curve, and the ideal curve has a high degree of fit.

### Model visualization

Based on the three clinical prediction models we developed to identify septic arthritis and transient synovitis in children, we built an easy-to-use online tool to predict the risk of septic arthritis and transient synovitis(http://www.mskca.tech/septicarthritis). It can calculate the probability of a child having septic arthritis or transient synovitis, thereby providing a more intuitive and understandable way to interpret the prediction model. This online tool helps doctors differentiate between septic arthritis and transient synovitis in children and choose the appropriate treatment (Figure 4).

**Figure 4.**
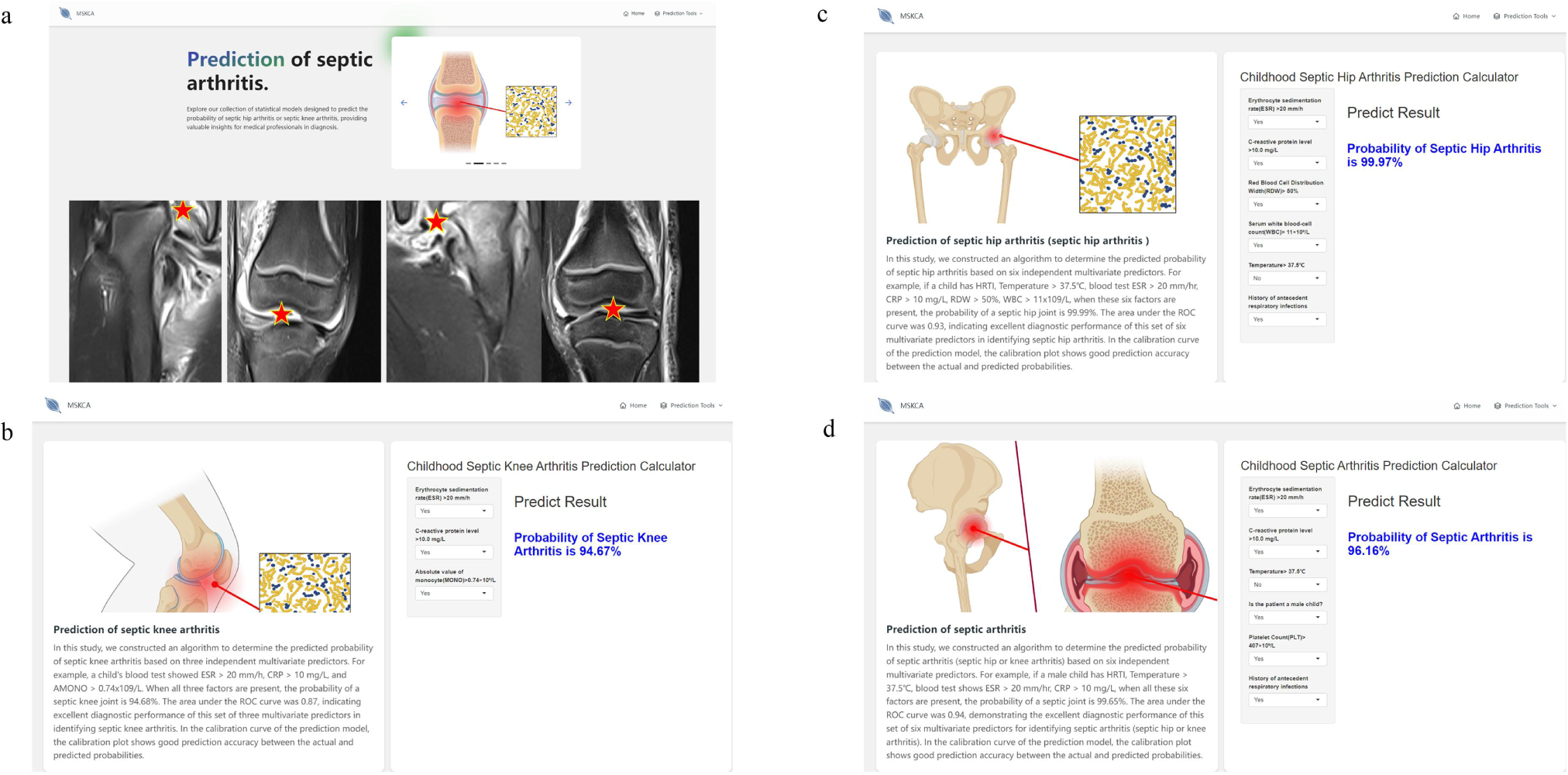
a. Main page of the online tool for predicting the risk of septic arthritis and transient synovitis. b. Sub-page of the online tool for predicting the risk of septic knee arthritis. c. Sub-page of the online tool for predicting the risk of septic hip arthritis. d. Sub-page of the online tool for predicting the risk of septic hip arthritis or septic knee arthritis.

## Discussion

Given the challenge of differentiating septic arthritis from transient synovitis in children, this study aimed to determine the diagnostic value for distinguishing these two conditions and to develop an effective clinical prediction model based on multi-center clinical data to identify and differentiate between the two diseases more comprehensively and accurately. We collected multiple-center data of 819 children from 8 tertiary specialized children’s hospitals through multiple centers, including 265 patients with septic arthritis and 554 patients with transient synovitis. Tand the mean age of the children with septic arthritis was 5.55±7.16 years, and the mean age of those with transient synovitis was 4.8±3.2 years. Among the blood and joint aspiration cultures of 265 cases of septic arthritis, the top 3 pathogenic bacteria were

Staphylococcus aureus (59%), Streptococcus pyogenes (10%), and Streptococci pneumoniae (7%). Furthermore, among the 265 patients with septic arthritis, 179 underwent surgical treatment; of them, 141 underwent incision and drainage + VSD negative pressure drainage of the affected joint, and 38 underwent arthroscopy-assisted knee joint lavage and drainage. To ensure the quality and reliability of the prediction model, we established three clinical prediction models:

- For septic hip arthritis, we used six clinical predictors: HRTI, temperature>37.5 ℃, ESR>20 mm/h, CRP>10 mg/L, RDW>50%, and WBC>11×10^9^/L. When all these six factors were present, the probability of having septic hip arthritis was 99.99%.
- For septic knee arthritis, we used three clinical predictors: ESR>20 mm/h, CRP>10 mg/L, and AMONO>0.74×10^9^/L. When all these three factors were present, the probability of having septic knee arthritis was 94.68%.
- For septic arthritis (septic hip arthritis or septic knee arthritis), we used six clinical predictors: male sex, HRTI, temperature>37.5 ℃, ESR>20 mm/h, CRP>10 mg/L, and PC>407×10^9^/L. When all these six factors were present, the probability of having septic arthritis was 99.65%.

Therefore, in addition to considering body temperature, peripheral WBC, ESR, and CRP, our clinical prediction model found new clinical predictors, including sex, HRTI, RDW, PC and AMONO.

### Analysis of pathogenic bacteria and empiric use of antibiotics

Our findings are consistent with worldwide research reports that Staphylococcus aureus is the most common causative bacterium for septic arthritis in children [10–12]. Moreover, in recent years, MRSA reports have gradually increased, and significant geographical differences exist in the prevalence of MRSA. MRSA was not found in patients with osteomyelitis in Saudi Arabia [13] or in bone and joint infections in Finland [14]; whereas in the United States, MRSA accounts for 30‒40% of bone and joint infections [15]. In our study, MRSA accounted for 12% of Staphylococcus aureus-linked cases. Streptococcus is reported to be the second most pathogenic microorganism in bone and joint infections, and Streptococcus infection causes disastrous consequences [16], in line with our findings. Children with bone and joint infections usually start empiric antibiotic treatment immediately after obtaining and submitting joint or bone specimens for examination. The selection of empiric treatment is to cover the most likely pathogens and is mainly based on the prevalence and resistance level of local pathogenic bacteria, the age of the child, and early laboratory results [17]. A Finnish study showed success as an initial empiric antibiotic therapy [14]. Therefore, understanding the epidemiological characteristics of pathogenic bacteria is crucial for developing effective treatment strategies in empiric antibiotic treatment against bone and joint infections in children.

### Septic arthritis (septic hip arthritis or septic knee arthritis) clinical prediction model

Septic arthritis and transient synovitis in children can sometimes be difficult to differentiate; however, accurate diagnosis is crucial for initiating different treatments and avoiding complications [18]. There is no simple test with high sensitivity, specificity, and ease of administration, to distinguish between the two conditions [19]. Clinicians consider various factors, including medical history, physical examination, and laboratory tests, when making a diagnosis. Clinical prediction models utilize evidence-based mathematical analysis to derive probabilities of diagnoses, helping clinicians to make objective decisions based on disease risk [20–22]. When faced with the critical but often difficult task of differentiating septic arthritis from transient synovitis in children, a probabilistic algorithm predicting septic arthritis based on independent multivariate predictors could be valuable in guiding diagnostic processes and ensuring timely, accurate diagnoses.

Previously, Kocher et al. used retrospective data to develop a clinical prediction algorithm to distinguish septic arthritis from transient synovitis. This algorithm uses four independent multivariate predictors and has excellent diagnostic performance in septic arthritis, with a 99.6% probability of diagnosis when all four factors are present [1]. However, Luhmann et al., who retrospectively applied the Kocher algorithm to determine its predictive value in a study of children, did not find the Kocher algorithm effective in their study, reporting a 59% probability of infectious arthritis when all four factors were present [5]. In 2006, Caird et al. used elevated CRP levels ≥20 mg/L as the fifth predictor, achieving a 98% probability of predicting septic arthritis [6].

The hip and knee joints are the most common joints affected by septic arthritis in children [8, 23]. To ensure the quality and reliability of the prediction model, we developed a model for septic hip arthritis based on six clinical predictors: HRTI, temperature>37.5 ℃, ESR>20 mm/h, CRP>10 mg/L, RDW>50%, and WBC>11×10^9^/L. The probability of septic hip arthritis was 99.99% when all six factors were present. We also developed a model for septic arthritis (septic hip arthritis or septic knee arthritis) based on six clinical predictors: male sex, HRTI, temperature>37.5 ℃, ESR>20 mm/h, CRP>10 mg/ L, and PC>407×10^9^/L. The probability of septic arthritis was 99.65% when all six factors were present.

Our clinical prediction model algorithm outperformed those from previous studies, with respect to the probability of diagnosing septic arthritis. In addition to considering body temperature, WBC, ESR, and CRP, we identified new clinical predictors, including sex, HRTI, PC and RDW. Consistent with previous reports, male children are more likely to develop septic arthritis than female children, with septic arthritis being 1.4–1.7 times more common in males than in females [24–27]. Septic arthritis is an infectious damage to the synovial fluid and joints caused by pathogenic microorganisms. The spread of infection through the blood to the joints is the most common transmission route in children after an upper respiratory tract infection; therefore, respiratory pathogens should be suspected as the cause of septic arthritis in pediatric patients [28, 29]. Similar to our findings, RDW is now recognized as a marker of systemic inflammation [30]. Most laboratories consider RDW values between 11.5% and 14.5% as normal. While traditionally used to differentiate between causes of anemia, an increased RDW is a marker of disrupted red blood cell homeostasis and is now thought to be an independent risk factor for systemic inflammation, critical illness morbidity, and mortality [31, 32]. Recent studies have suggested that increased RDW is associated with inflammatory biomarkers and oxidative stress [33, 34]; some prospective and retrospective studies have shown an abnormal increase in RDW to be an independent risk factor for death in patients with sepsis and septic shock [35].

Consistent with our findings, a reactive PC is frequently observed in children with infections or other causes of acute-phase reactions[36]. Studies have reported that increased PC may be a marker of the severity of lower respiratory tract infections in pediatric patients[37]. Likewise, in addition, elevated PC may have diagnostic and prognostic roles in the management of pediatric urinary tract infections, and results from a study of children with urinary tract infections suggest that elevated PC may be a symptom of upper urinary tract infections and Gram-positive Bacterial signs[38].

### Clinical prediction model for septic knee arthritis

While several studies have focused on developing clinical prediction models for septic hip arthritis and transient hip synovitis in children, few have investigated the diagnostic gold standard and related models for septic knee arthritis. An early study attempted to diagnose inflammation in children’s knee joint infections by applying the four clinical predictors proposed by Kocher et al.[1] but found that these factors did not apply to knee joint infections [9]. In contrast, another study found that the clinical prediction model algorithm proposed by Caird performed better than Kocher’s algorithm and that the Caird algorithm was clinically reasonable for children’s large joints, such as knee, ankle, shoulder, elbow, and wrist joints [39]. In our study, we used three clinical predictors for septic knee arthritis: ESR>20 mm/h, CRP>10 mg/L, and AMONO>0.74×10^9^/L. The probability of a patient being diagnosed with septic knee arthritis was 94.68%. Besides ESR and CRP, we introduced AMONO as a new diagnostic indicator for septic knee arthritis. Similar to our research results, monocytes have been reported to play an important role in inflammation. Furthermore, common causes of increased monocyte count include infection, leukemia, and autoimmune diseases [40], which support our view of monocytes as a new factor in septic knee arthritis diagnosis. This finding further strengthens our proposition of AMONO as a valuable tool for diagnosing septic arthritis.

## Conclusion

This study developed a new clinical prediction model for septic arthritis in children using multi-center data. Our data support the high application value of this model in clinical practice, providing more accurate diagnosis and prediction of septic arthritis in children. These findings and recommendations can increase the vigilance of frontline physicians and help them to prepare for the initial evaluation and management of pediatric patients with suspected septic arthritis, minimizing painful procedures and unnecessary treatments and optimizing patient outcomes.

### Limitations

This study has some limitation. Data were collected from eight pediatric tertiary specialty hospitals in China. This may have led to inconsistencies in case records and differences in clinical assessments. In addition, this was a retrospective study, therefore, it is not possible to control potential confounding factors as well as the impact of selection bias, and future studies should include a prospective evaluation of the diagnostic performance of the clinical prediction algorithm at the same site, in addition to prospective validations in new clinical settings.

## Authors contribution

Prof. Zhu Xiong, Prof. Sheng-Ping Tang and Prof. Qi-Ru Su supervised and guided the project, wrote and revised the manuscript. Dr.Xin Qiu, Dr.Han-Sheng Deng and Dr.Gen Tang collected the data. Dr. Xiao-Liang Chen, Dr. Yao-Xi Liu, Dr. Jing-Chun Li, Dr. Xin-Wu Wu, Dr. Jia-Chao Guo and Dr. Fei Jiang provided and analysed the clinical data and guided the research.

## Availability of data

The data sets used and analysed during the current study are available from these corresponding authors on reasonable request.

## Supporting information

none

none

none

none

none

none

none

## Data Availability

http://www.mskca.tech/septicarthritis

## Acknowledgements

We would like to express gratitude to all the patients and their families.

## References

1. Kocher, M.S., D. Zurakowski, and J.R. Kasser, Differentiating between septic arthritis and transient synovitis of the hip in children: an evidence-based clinical prediction algorithm. J Bone Joint Surg Am, 1999. 81(12): p. 1662–70.

2. Sultan, J. and P.J. Hughes, Septic arthritis or transient synovitis of the hip in children: the value of clinical prediction algorithms. J Bone Joint Surg Br, 2010. 92(9): p. 1289–93.

3. Dan, M., Septic arthritis in young infants: clinical and microbiologic correlations and therapeutic implications. Rev Infect Dis, 1984. 6(2): p. 147–55.

4. Haueisen, D.C., D.S. Weiner, and S.D. Weiner, The characterization of “transient synovitis of the hip” in children. J Pediatr Orthop, 1986. 6(1): p. 11–7.

5. Luhmann, S.J., et al., Differentiation between septic arthritis and transient synovitis of the hip in children with clinical prediction algorithms. J Bone Joint Surg Am, 2004. 86(5): p. 956–62.

6. Caird, M.S., et al., Factors distinguishing septic arthritis from transient synovitis of the hip in children. A prospective study. J Bone Joint Surg Am, 2006. 88(6): p. 1251–7.

7. Sakwe, N., et al., Relationship between malaria, anaemia, nutritional and socio-economic status amongst under-ten children, in the North Region of Cameroon: A cross-sectional assessment. PLoS One, 2019. 14(6): p. e0218442.

8. Al Saadi, M.M., et al., Acute septic arthritis in children. Pediatr Int, 2009. 51(3): p. 377–80.

9. Joshy, S., et al., Comparison of bacteriologically proven septic arthritis of the hip and knee in children, a preliminary study. J Pediatr Orthop, 2010. 30(2): p. 208–11.

10. Kang, S.N., et al., The management of septic arthritis in children: systematic review of the English language literature. J Bone Joint Surg Br, 2009. 91(9): p. 1127–33.

11. Basmaci, R., et al., Comparison of clinical and biologic features of Kingella kingae and Staphylococcus aureus arthritis at initial evaluation. Pediatr Infect Dis J, 2011. 30(10): p. 902–4.

12. Cohen, E., et al., Septic arthritis in children: Updated epidemiologic, microbiologic, clinical and therapeutic correlations. Pediatr Neonatol, 2020. 61(3): p. 325–330.

13. Kopp, V.J.J.P.I.D., The clinical profile of childhood osteomyelitis: A Saudi experience. 2008. 03(04): p. 235–240.

14. Peltola, H., et al., Clindamycin vs. first-generation cephalosporins for acute osteoarticular infections of childhood--a prospective quasi-randomized controlled trial. Clin Microbiol Infect, 2012. 18(6): p. 582–9.

15. Arnold, S.R., et al., Changing patterns of acute hematogenous osteomyelitis and septic arthritis: emergence of community-associated methicillin-resistant Staphylococcus aureus. J Pediatr Orthop, 2006. 26(6): p. 703–8.

16. Kuong, E.E., et al., Pitfalls in diagnosing septic arthritis in Hong Kong children: ten years’ experience. Hong Kong Med J, 2012. 18(6): p. 482–7.

17. He, M., et al., An update on recent progress of the epidemiology, etiology, diagnosis, and treatment of acute septic arthritis: a review. Front Cell Infect Microbiol, 2023. 13: p. 1193645.

18. Donders, C.M., et al., Developments in diagnosis and treatment of paediatric septic arthritis. World J Orthop, 2022. 13(2): p. 122–130.

19. Donders, C.M., et al., Arthrocentesis, arthroscopy or arthrotomy for septic knee arthritis in children: a systematic review. J Child Orthop, 2021. 15(1): p. 48–54.

20. Ranstam, J., J.A. Cook, and G.S. Collins, Clinical prediction models. Br J Surg, 2016. 103(13): p. 1886.

21. Collins, G.S., et al., Transparent Reporting of a multivariable prediction model for Individual Prognosis or Diagnosis (TRIPOD): the TRIPOD statement. Ann Intern Med, 2015. 162(1): p. 55–63.

22. Moons, K.G.M., et al., Prognosis and prognostic research: What, why, and how? 2009. 338(feb23 1): p. b375.

23. Rutz, E. and R. Brunner, Septic arthritis of the hip - current concepts. Hip Int, 2009. 19 Suppl 6: p. S9–12.

24. Riise Ø, R., et al., Incidence and characteristics of arthritis in Norwegian children: a population-based study. Pediatrics, 2008. 121(2): p. e299–306.

25. Pääkkönen, M., Septic arthritis in children: diagnosis and treatment. Pediatric Health Med Ther, 2017. 8: p. 65–68.

26. Okubo, Y., K. Nochioka, and T. Marcia, Nationwide survey of pediatric septic arthritis in the United States. J Orthop, 2017. 14(3): p. 342–346.

27. Peltola, H., et al., Prospective, randomized trial of 10 days versus 30 days of antimicrobial treatment, including a short-term course of parenteral therapy, for childhood septic arthritis. Clin Infect Dis, 2009. 48(9): p. 1201–10.

28. Martin, J., M.D. Morlius, and M.A.J.R.C.E. Falgueras, Septic arthritis due to Streptococcus pneumoniae. Report of two cases of unusual location. 1998. 198(9): p. 596–597.

29. Mooney, J.F., 3rd and R.F. Murphy, Septic arthritis of the pediatric hip: update on diagnosis and treatment. Curr Opin Pediatr, 2019. 31(1): p. 79–85.

30. Salvagno, G.L., et al., Red blood cell distribution width: A simple parameter with multiple clinical applications. Crit Rev Clin Lab Sci, 2015. 52(2): p. 86–105.

31. Viallat, A. and M. Abkarian, Red blood cell: from its mechanics to its motion in shear flow. Int J Lab Hematol, 2014. 36(3): p. 237–43.

32. Bateman, R.M., et al., The Effect of Sepsis on the Erythrocyte. Int J Mol Sci, 2017. 18(9).

33. Bulut, O., et al., Elevated Red Cell Distribution Width as a Useful Marker in Neonatal Sepsis. J Pediatr Hematol Oncol, 2021. 43(5): p. 180–185.

34. Mohanty, J.G., E. Nagababu, and J.M. Rifkind, Red blood cell oxidative stress impairs oxygen delivery and induces red blood cell aging. Front Physiol, 2014. 5: p. 84.

35. Sadaka, F., J. O’Brien, and S. Prakash, Red cell distribution width and outcome in patients with septic shock. J Intensive Care Med, 2013. 28(5): p. 307–13.

36. Cecinati, V., L. Brescia, and S. Esposito, Thrombocytosis and infections in childhood. Pediatr Infect Dis J, 2012. 31(1): p. 80–1.

37. Vlacha, V. and G. Feketea, Thrombocytosis in pediatric patients is associated with severe lower respiratory tract inflammation. Arch Med Res, 2006. 37(6): p. 755–9.

38. Garoufi, A., et al., Reactive thrombocytosis in children with upper urinary tract infections. Acta Paediatr, 2001. 90(4): p. 448–9.

39. Nickel, A.J., et al., Novel Uses of Traditional Algorithms for Septic Arthritis. J Pediatr Orthop, 2022. 42(2): p. e212–e217.

40. Cren, M., et al., Differential Accumulation and Activation of Monocyte and Dendritic Cell Subsets in Inflamed Synovial Fluid Discriminates Between Juvenile Idiopathic Arthritis and Septic Arthritis. Front Immunol, 2020. 11: p. 1716.

