## Supplementary material for "Clinical prediction model performance in differentiating septic arthritis from transient synovitis: A multi-center study": none

Table1.Univariate logistic regression analysis of septic hip arthritis and transient hip synovitis

| Variable | Regression Coefficient | Odds Ratio | Ninety-five Percent Confidence Interva | P Value |
| --- | --- | --- | --- | --- |
| Erythrocyte sedimentation rate | 0.07 | 1.07 | 1.05-1.09 | <0.01 |
| Peripheral blood erythrocyte count | -3.20 | 0.04 | 0.02-0.08 | <0.01 |
| Hemoglobin | -0.10 | 0.90 | 0.89-0.92 | <0.01 |
| C-reactive protein | 0.09 | 1.09 | 1.07-1.12 | <0.01 |
| Red blood cell distribution width | -0.13 | 0.88 | 0.86-0.9 | <0.01 |
| History of preceding respiratory tract infection | 1.93 | 6.89 | 4.39-10.81 | <0.01 |
| Absolute monocyte count | 2.92 | 18.54 | 8.8-39.05 | <0.01 |
| Hematocrit | -0.32 | 0.73 | 0.67-0.79 | <0.01 |
| Eosinophil percentage | -0.68 | 0.51 | 0.42-0.6 | <0.01 |
| Lymphocyte percentage | -0.06 | 0.94 | 0.92-0.96 | <0.01 |
| Serum white blood cell | 0.23 | 1.26 | 1.19-1.33 | <0.01 |
| Initial hospital admission temperature | 1.47 | 4.35 | 2.83-6.69 | <0.01 |
| Absolute neutrophil count | 0.24 | 1.27 | 1.18-1.37 | <0.01 |
| Platelet Count (PC) | 0.01 | 1.01 | 1.01-1.01 | <0.01 |
| Platelet crit | 6.49 | 658.52 | 89.19-4861.97 | <0.01 |
| Mean corpuscular hemoglobin concentration | -0.05 | 0.95 | 0.93-0.97 | <0.01 |
| Mean corpuscular volume | 0.10 | 1.11 | 1.06-1.15 | <0.01 |
| Absolute eosinophil count | -3.14 | 0.04 | 0.01-0.17 | <0.01 |
| Male | -1.00 | 0.37 | 0.24-0.57 | <0.01 |
| Platelet distribution width | 0.19 | 1.21 | 1.12-1.31 | <0.01 |
| Neutrophil percentage | -0.02 | 0.98 | 0.96-1 | <0.01 |
| Age | -0.15 | 0.86 | 0.8-0.93 | <0.01 |
| Absolute lymphocyte count | 0.23 | 1.26 | 1.12-1.42 | <0.01 |
| Monocyte proportion | 0.09 | 1.09 | 1.01-1.18 | 0.01 |
| Mean corpuscular hemoglobin content | 0.08 | 1.08 | 0.98-1.19 | 0.12 |
| History of prodromal strenuous exercise | -0.41 | 0.66 | 0.29-1.54 | 0.34 |
| Mean platelet volume (MPV) | 0.01 | 1.01 | 0.83-1.23 | 0.90 |
