## Supplementary material for "Clinical prediction model performance in differentiating septic arthritis from transient synovitis: A multi-center study": none

Table2.Univariate logistic regression analysis of septic knee arthritis and transient knee synovitis

| Variable | Regression Coefficient | Odds Ratio | Ninety-five Percent Confidence Interva | P Value |
| --- | --- | --- | --- | --- |
| Erythrocyte sedimentation rate | 0.06 | 1.06 | 1.04-1.09 | <0.01 |
| C-reactive protein | 0.12 | 1.13 | 1.07-1.19 | <0.01 |
| Absolute monocyte count | 2.51 | 12.24 | 3.18-47.16 | <0.01 |
| Eosinophil percentage | -0.35 | 0.70 | 0.58-0.85 | <0.01 |
| Initial hospital admission temperature | 1.39 | 4.01 | 1.77-9.08 | <0.01 |
| Hemoglobin | -0.05 | 0.95 | 0.93-0.98 | <0.01 |
| Serum white blood cell | 0.17 | 1.18 | 1.07-1.32 | <0.01 |
| Peripheral blood erythrocyte count | -1.32 | 0.27 | 0.12-0.62 | <0.01 |
| Platelet Count (PC) | 0.01 | 1.01 | 1-1.01 | <0.01 |
| Hematocrit | -0.16 | 0.85 | 0.76-0.95 | <0.01 |
| Platelet crit | 4.88 | 130.97 | 4.64-3695.38 | <0.01 |
| Absolute neutrophil count | 0.20 | 1.22 | 1.06-1.4 | 0.01 |
| Absolute eosinophil count | -2.60 | 0.07 | 0.01-0.49 | 0.01 |
| Mean corpuscular hemoglobin concentration | -0.03 | 0.97 | 0.94-1 | 0.04 |
| Lymphocyte percentage | -0.01 | 0.99 | 0.97-1 | 0.15 |
| Monocyte proportion | 0.06 | 1.06 | 0.97-1.17 | 0.20 |
| Absolute lymphocyte count | 0.11 | 1.11 | 0.93-1.32 | 0.23 |
| History of preceding respiratory tract infection | 0.38 | 1.46 | 0.67-3.16 | 0.34 |
| Neutrophil percentage | 0.01 | 1.01 | 0.99-1.02 | 0.35 |
| Age | -0.04 | 0.96 | 0.87-1.06 | 0.41 |
| History of prodromal strenuous exercise | -0.41 | 0.66 | 0.22-1.99 | 0.46 |
| Mean platelet volume (MPV) | -0.11 | 0.90 | 0.64-1.26 | 0.54 |
| Red blood cell distribution width | -0.01 | 0.99 | 0.96-1.02 | 0.57 |
| Mean corpuscular volume | 0.02 | 1.02 | 0.96-1.08 | 0.58 |
| Mean corpuscular hemoglobin content | -0.04 | 0.96 | 0.81-1.13 | 0.61 |
| Mean corpuscular hemoglobin concentration | 0.02 | 1.02 | 0.93-1.13 | 0.65 |
| Male | -0.003 | 0.997 | 0.48-2.08 | 0.99 |
