## Supplementary material for "Clinical prediction model performance in differentiating septic arthritis from transient synovitis: A multi-center study": none

Table3. Univariate logistic regression analysis of septic arthritis (septic hip arthritis or septic knee arthritis) and transient synovitis (transient hip synovitis or transient knee synovitis)

| Variable | Regression Coefficient | Odds Ratio | Ninety-five Percent Confidence Interva | P Value |
| --- | --- | --- | --- | --- |
| Erythrocyte sedimentation rate | 0.07 | 1.07 | 1.06-1.09 | <0.01 |
| C-reactive protein | 0.09 | 1.10 | 1.07-1.12 | <0.01 |
| Absolute monocyte count | 2.92 | 18.59 | 8.87-38.97 | <0.01 |
| Eosinophil percentage | -0.68 | 0.51 | 0.42-0.61 | <0.01 |
| Initial hospital admission temperature | 1.47 | 4.35 | 2.84-6.66 | <0.01 |
| Hemoglobin | -0.10 | 0.90 | 0.88-0.92 | <0.01 |
| Serum white blood cell | 0.23 | 1.25 | 1.18-1.33 | <0.01 |
| Peripheral blood erythrocyte count | -3.20 | 0.04 | 0.02-0.08 | <0.01 |
| Platelet Count (PC) | 0.01 | 1.01 | 1-1.01 | <0.01 |
| Hematocrit | -0.32 | 0.73 | 0.67-0.79 | <0.01 |
| Platelet crit | 6.49 | 659.27 | 89.04-4881.29 | <0.01 |
| Absolute neutrophil count | 0.24 | 1.27 | 1.18-1.36 | <0.01 |
| Absolute eosinophil count | -3.14 | 0.04 | 0.01-0.17 | <0.01 |
| Mean corpuscular hemoglobin concentration | -0.05 | 0.95 | 0.93-0.97 | <0.01 |
| Lymphocyte percentage | -0.06 | 0.94 | 0.93-0.96 | <0.01 |
| Absolute lymphocyte count | 0.23 | 1.25 | 1.11-1.42 | <0.01 |
| History of preceding respiratory tract infection | 1.93 | 6.86 | 4.34-10.84 | <0.01 |
| Neutrophil percentage | -0.02 | 0.98 | 0.97-0.99 | <0.01 |
| Age | -0.15 | 0.86 | 0.79-0.93 | <0.01 |
| Red blood cell distribution width | -0.13 | 0.88 | 0.86-0.91 | <0.01 |
| Mean corpuscular volume | 0.10 | 1.10 | 1.06-1.15 | <0.01 |
| Platelet distribution width | 0.19 | 1.21 | 1.11-1.32 | <0.01 |
| Male | -1.001 | 0.37 | 0.24-0.57 | <0.01 |
| Monocyte proportion | 0.09 | 1.10 | 1.02-1.18 | 0.01 |
| History of prodromal strenuous exercise | -0.41 | 0.66 | 0.28-1.53 | 0.34 |
| Mean platelet volume | 0.01 | 1.01 | 0.83-1.24 | 0.90 |
| Mean corpuscular hemoglobin content | 0.08 | 1.08 | 0.98-1.2 | 0.12 |
