## Supplementary material for "Clinical prediction model performance in differentiating septic arthritis from transient synovitis: A multi-center study": none

Table4.Multivariate analysis of septic hip arthritis and transient hip synovitis

| Variable | Regression Coefficient | Likehood Ratio | P Value Of Likehood Ratio Test | Adjusted Odds Ratio | Ninety-five Percent Confidence Interva | P Value |
| --- | --- | --- | --- | --- | --- | --- |
| ESR>20mm/hr | 2.01 | 33.99 | <0.01 | 7.43 | 3.7-14.92 | <0.01 |
| CRP>10mg/L | 2.51 | 59.36 | <0.01 | 12.30 | 6.3-24 | <0.01 |
| RDW>50% | 2.92 | 6.77 | 0.01 | 18.62 | 1.29-268.44 | 0.03 |
| WBC>11×10^9^/L | 0.68 | 3.91 | 0.05 | 1.98 | 1.01-3.89 | 0.05 |
| Temperature>37.5℃ | 2.32 | 16.24 | <0.01 | 10.20 | 3.12-33.38 | <0.01 |
| HRTI (YES) | 1.45 | 17.40 | <0.01 | 4.28 | 2.15-8.49 | <0.01 |
