## Supplementary material for "Clinical prediction model performance in differentiating septic arthritis from transient synovitis: A multi-center study": none

Table5.Multivariate analysis of septic knee arthritis and transient knee synovitis

| Variable | Regression Coefficient | Likehood Ratio | P Value Of Likehood Ratio Test | Adjusted Odds Ratio | Ninety-five Percent Confidence Interva | P Value |
| --- | --- | --- | --- | --- | --- | --- |
| ESR>20mm/hr | 1.51 | 8.12 | <0.01 | 4.54 | 1.6-12.89 | <0.01 |
| CRP>10mg/L | 1.80 | 10.63 | <0.01 | 6.05 | 2.02-18.13 | <0.01 |
| AMONO>0.74×10^9^/L | 1.23 | 5.48 | 0.02 | 3.44 | 1.18-10.03 | 0.02 |
