## Supplementary material for "Clinical prediction model performance in differentiating septic arthritis from transient synovitis: A multi-center study": none

Table6. Multivariate analysis of septic arthritis (septic hip arthritis or septic knee arthritis) and transient synovitis(transient hip synovitis or transient knee synovitis)

| Variable | Regression Coefficient | Likehood Ratio | P Value Of Likehood Ratio Test | Adjusted Odds Ratio | Ninety-five Percent Confidence Interva | P Value |
| --- | --- | --- | --- | --- | --- | --- |
| ESR>20mm/hr | 1.90 | 43.96 | <0.01 | 6.65 | 3.75-11.8 | <0.01 |
| CRP>10mg/L | 2.52 | 83.01 | <0.01 | 12.47 | 7.02-22.17 | <0.01 |
| Temperature>37.5℃ | 2.42 | 28.26 | <0.01 | 11.28 | 4.32-29.42 | <0.01 |
| Male gender(YES) | -0.76 | 6.93 | 0.01 | 0.47 | 0.27-0.83 | 0.01 |
| PC>407×10^9^/L | 2.05 | 47.55 | <0.01 | 7.74 | 4.18-14.31 | <0.01 |
| HRTI (YES) | 0.69 | 5.44 | 0.02 | 1.99 | 1.12-3.55 | 0.02 |
